# Comparing brain structural effects of dopaminergic antagonism and partial agonism in antipsychotic-naïve patients with first-episode psychosis using normative modeling

**DOI:** 10.1101/2025.07.30.25332427

**Authors:** Anna Feveile, Karen S. Ambrosen, Ketevan Shalikashvili, Jayachandra M. Raghava, Mette Ø. Nielsen, Kirsten B. Bojesen, Barbora R. Bučková, Andre F. Marquand, Birte Y. Glenthøj, Warda T. Syeda, Bjørn H. Ebdrup

## Abstract

**Aim:** Schizophrenia is associated with subtle brain structural alterations but separating disease from medication effects is challenging. Antipsychotic dopamine D2 receptor (D2R) antagonism has been associated with striatal volume increases, but effects of partial D2R agonism by newer antipsychotics are largely unexplored. This study aimed to compare short-term brain changes associated with either D2R antagonism or partial D2R agonism using normative modeling. Secondarily, the study aimed to explore long-term effects following naturalistic treatment.

**Methods:** Patients received 6 weeks of monotherapy with either amisulpride (D2R antagonist) (N=41) or aripiprazole (partial D2R agonist) (N=45). All patients underwent structural magnetic resonance imaging before and after 6 weeks of treatment. A subset was re-scanned at 6 months, 1 year, and 2 years of naturalistic treatment. A pre-trained normative model was applied to 186 subcortical and cortical regions. We used Wilcoxon signed-rank tests to identify longitudinal structural deviations.

**Results:** Amisulpride and aripiprazole were associated with striatal volume increases after six weeks. No cortical effects were observed with amisulpride. Thinning of the temporal lobe was observed with aripiprazole. After 6 months, and 1 and 2 years, the striatal changes abated but cortical thinning in the frontal lobes emerged.

**Conclusions:** Both partial D2R agonism and D2R antagonism appear linked to striatal volume increases, however changes appear transient. Conversely, frontal thinning occurs over time and appears less closely linked to antipsychotic treatment. Interpretating structural brain changes in patients with psychosis require consideration of short-term pharmacological effects as well as factors related to illness progression.

## Introduction

Schizophrenia is a severe psychiatric disorder that affects around 1% of the population (1). Patients with more chronic states of schizophrenia have been shown to display progressive cortical thinning in the frontal and temporal lobes (2,3). Subcortical structures, including the hippocampus, amygdala, and thalamus are already affected in the earlier phases of psychosis (4). However, patients display high brain structural heterogeneity (5), and reports of affected brain regions differ between studies and patient samples (6). The pathophysiology of schizophrenia has been linked to presynaptic dysregulation in the dopaminergic pathways (7), and modulation of dopamine receptors is the key approach to pharmacological treatment with antipsychotics (APs). All currently licensed APs target dopamine D2 receptors (D2Rs), and AP-mediated striatal D2R blockade has consistently been linked to treatment response (8).

Multiple studies have suggested that long-term exposure to higher doses of AP is associated with more extensive changes to brain morphology (9–11). Previous studies on rodents have identified striatal volume increases following treatment with APs blocking D2Rs (12–14). Similarly, longitudinal MRI studies on initially AP-naïve patients reported striatal volume increases after three months of AP monotherapy (15,16). By means of conventional volumetric analyses on structural MRI data from one of the cohorts included in the present dataset, we previously reported increases in the bilateral caudate nucleus and right putamen after only six weeks of monotherapy with the second-generation AP, amisulpride (17).

Antipsychotics have traditionally been categorized into first- or second-generation. However, this classification was challenged by the approval in 2002 of aripiprazole which acts on D2Rs through partial agonism (18). In 2015, two additional partial D2R agonists, brexpiprazole and cariprazine were approved. From a pharmacological perspective these partial agonists can be considered ‘third generation antipsychotics”. Third generation APs are largely recommended as first-line treatment for psychosis due to their generally more favorable side-effect profile (19). We have recently shown that six weeks exposure to aripiprazole increases the resting striatal blood flow (20). Moreover, the increase in striatal perfusion seem to normalize after two years (21). However, the volumetric correlates to these observations have not been investigated. Since some of the initial studies documenting brain structural effects of APs sparked major scientific and public concern (e.g. (22)), it is pertinent to investigate the brain structural effects of third-generation APs.

Due to the heterogeneous presentation of schizophrenia, innovative statistical methods are needed to improve our interferences about the effects of treatment. Normative modeling has emerged as a powerful tool for characterizing individual-level deviations in brain structure. This approach, akin to pediatric growth charts, allows researchers to map deviations from expected neurodevelopmental trajectories, thereby capturing the heterogeneity often observed in psychiatric populations (23,24). It has been widely applied to imaging-derived phenotypes (IDPs) in patients with various neurodevelopmental and psychiatric conditions (25–29) and can be used to track longitudinal changes relative to age-normative baselines (30).

Here, we applied a pre-trained normative model (30,31) to measures of cortical thickness and subcortical volume from AP-naïve patients with first-episode psychosis (FEP) to investigate short-term brain structural changes after six weeks of AP monotherapy with either a D2R antagonist, amisulpride, or a partial D2R agonist, aripiprazole. Furthermore, we investigated the more long-term structural changes after 6 months, 1 year, and 2 years of naturalistic treatment.

Using a normative modeling approach, we first aimed to confirm previous conventional analyses showing volumetric striatal increases after six weeks of amisulpride treatment. Second, we hypothesized that patients treated with aripiprazole for six weeks would display less pronounced volumetric increases in the striatum compared to patients treated with amisulpride. Finally, we expected that patients at long-term follow-up timepoints would conform to a more chronic stage of psychosis by displaying cortical thinning of the frontal and temporal lobes (2,3).

## Methods

### Participants

Antipsychotic-naïve patients with first-episode psychosis were recruited from in- and out-patient clinics in the Capital Region of Denmark as part of the two consecutive observational cohort studies: Pan European Collaboration on Antipsychotic Naïve Schizophrenia (PECANS; ClinicalTrials.gov identifier: NCT01154829; 2008-2016) (32) and Pan European Collaboration on Antipsychotic Naïve Schizophrenia II (PECANSII; ClinicalTrials.gov identifier: NCT02339844; 2014-2021) (33). The two studies were approved by the Regional Danish Committee on Health Research Ethics (H-D-2008-088 and H-3-2013-149) and conducted in accordance with the Declaration of Helsinki. All participants provided written informed consent.

Patients were between 18 and 45 years of age and met the International Classification of Diseases (ICD-10) criteria for schizophrenia (F20.x, both cohorts), schizoaffective disorder (F25.x, PECANSII), or non-organic psychosis (F22.x, F28.x, F29.x; PECANSII). Diagnoses were verified by the Schedule of Clinical Assessment in Neuropsychiatry (SCAN) and confirmed by a psychiatrist. The exclusion criteria for patients included prior treatment with APs, treatment with antidepressants within the last month prior to baseline examination, current substance abuse, pregnancy, a history of severe medical conditions, or more than 5 minutes of unconsciousness. Healthy controls were recruited through advertisement and matched to patients on age, sex, and parental socioeconomic status. Exclusion criteria for healthy controls included psychiatric illness or psychiatric diagnoses in first-degree relatives, in addition to the exclusion criteria also applied to patients. All participants were subjected to urine tests to assess recent drug use (Rapid Response, Jepsen HealthCare, Tune, DK).

### Medication

All patients were AP-naïve at baseline. From baseline to six-week follow-up visit, the patients received AP monotherapy with either a D2R antagonist, amisulpride (PECANS, here referred to as “Amisulpride cohort”), or a partial D2R agonist, aripiprazole (PECANSII, here referred to as “Aripiprazole cohort”). Patients who did not receive AP treatment during the full six weeks were excluded from follow-up analyses. To ensure dose comparability across compounds, amisulpride and aripiprazole doses were converted to olanzapine dose equivalents using the weighted means method (34). After the six-week follow-up visit, the patients received naturalistic AP treatment.

### Clinical and cognitive measures

Symptom severity was assessed at all visits using the Positive and Negative Syndrome Scale (PANSS) (35), and subscale scores (PANSS_positive_, PANSS_negative_, and PANSS_general_) were calculated. To describe the change in PANSS scores from baseline to follow-up, we calculated the absolute change resulting in ΔPANSS_positive_, ΔPANSS_negative_, and ΔPANSS_general_, where ΔPANSS < 0 reflects a decrease in PANSS and thus improvement in symptoms. At baseline, the patients’ level of functioning was quantified using the Global Assessment of Functioning (GAF). IQ was estimated using the Wechsler Adult Intelligence Scale v. 3 (WAIS-III) (36), and handedness was assessed using the Edinburgh Handedness Inventory (EHI) (37).

### Magnetic resonance imaging and processing

T1-weighted MRI scans were obtained from all participants using a single 3T Philips Achieva MRI scanner (Philips Healthcare, Best, the Netherlands) with an 8-channel (Amisulpride cohort) or a 32-channel (Aripiprazole cohort) SENSE head coil (Invivo Corporation) at baseline and after 6 weeks, 6 months, 1 year, and 2 years. Scans were acquired using the parameters: TR = 10 ms, TE = 4.6 ms, FA = 8°, and voxel size = 0.79 mm × 0.79 mm × 0.80 mm. Images were processed in FreeSurfer v. 5.3.0 (38,39) using the longitudinal processing stream (40). A total of 186 imaging derived phenotypes (IDPs) describing regional cortical thickness (150 IDPs), based on the Destrieux cortical atlas (41), and regional subcortical volumes, based on a probabilistic atlas (36 IDPs) (42), were extracted for all scans. Image and FreeSurfer segmentation quality was assessed manually and using Qoala-T (43) separately for the two cohorts.

### Normative modeling

We applied a pre-trained normative model, available from PCNtoolkit (https://github.com/predictive-clinical-neuroscience/PCNtoolkit-demo/blob/main/tutorials/BLR_protocol/transfer_pretrained_normative_models.ipynb), run in Python v. 3.10.5 (Visual Studio Code v. 1.97.2). The model is a Bayesian linear regression model trained on 186 cortical thickness and regional volume IDPs (identical to those we extracted from FreeSurfer) from 58,836 healthy individuals, aged 2-100 years, collected from 82 different sites (31). For details on the model and its implementation, please, refer to papers by Fraza *et al.* (44) and Rutherford *et al.* (31), respectively. The normative model was adapted to our cohorts using data from approx. 40% of the healthy controls (HCs) (i.e., 47 HCs), stratified on age, sex, and cohort. This percentage was chosen to balance accurate adaptation with the retention of a sufficient number of healthy controls for subsequent analyses.

A successfully adapted normative model provides an individualized deviation score (Z-score), that reflect one’s position relative to the healthy population distribution.

Recently, this pre-trained model was adapted to longitudinally processed data by Bučková *et al.* (30), who introduced a trajectory deviation score (Z_diff_), describing the longitudinal change between two visits normalized to the healthy control group. Z_diff_=0 means that the IDP remains on the predicted trajectory, while Z_diff_>0 and Z_diff_<0 means that the IDP increases and decreases, respectively, relative to the predicted trajectory. The script used for longitudinal model application is available from PCNtoolkit (https://github.com/predictive-clinical-neuroscience/PCNtoolkit-demo/blob/main/tutorials/Long_NM/Long_NM_protocol.ipynb). The longitudinal modeling outputs are the patient- and IDP-specific trajectory deviation scores from baseline to each of the four follow-ups.

For more details on Z- and Z_diff_ scores, please refer to Supplementary Material.

### Statistical analyses

All statistical analyses were conducted in Python v. 3.10.5 (Visual Studio Code v. 1.97.2). The level of significance was defined as *⍺* = 0.05. Demographic and clinical differences between patients and controls were evaluated using two-sample t-test, Mann-Whitney U-test, Chi-squared test, or Fisher’s exact test as appropriate. Normality of continuous variables were evaluated by Shapiro-Wilk tests and inspection of histograms. Similarly, comparisons were conducted between patients from each cohort, and finally between patients with and without 6-week follow-up data to address attrition bias. To describe initial structural group differences, baseline Z-scores of patients and HCs were compared using mass Welch’s t-tests. To control the false discovery rate (FDR) at α = 0.05 for all 186 comparisons, we applied the Benjamini-Hochberg procedure (45).

Group-level longitudinal deviations from the predicted trajectory were determined by conducting mass Wilcoxon signed-rank tests on Z_diff_ scores from baseline to each follow-up. We report the group average 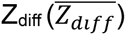 scores for significant IDPs to reflect the direction and magnitude of longitudinal deviations. First, longitudinal deviations were assessed using 6-week follow-up Z_diff_ scores from the Amisulpride and Aripiprazole cohort separately. In the long-term analyses, the two cohorts were pooled, and all participants with MRI data from baseline and the follow-up visit concerned were included. For each analysis, the p-values were subjected to the Benjamini-Hochberg procedure (45) to control the false discovery rate at α = 0.05 for the 186 IDPs.

For the structural analyses, we only report significant findings after correction for multiplicity, unless otherwise stated (as ‘p_uncorrected_’).

To address potential biases of the structural changes after six weeks, we conducted post-hoc analyses exploring correlations between Z_diff_ scores and change in body weight and AP dose within each cohort using Spearman’s rank-order correlation. Finally, we explored correlations between structural changes at all follow-ups and treatment response using both a univariate and a pattern-based approach using principal component analysis (PCA) (see Supplementary Material for details).

## Results

### Demographic and clinical characteristics

In the main analysis, we included 151 participants (86 AP-naïve patients with FEP and 65 HCs) with MRI data from baseline and six-week follow-up. Forty-one patients received amisulpride monotherapy and 45 patients received aripiprazole monotherapy. For an overview of subjects included in the analyses, please refer to Figure 1. At baseline, the duration of education was shorter (p<0.001) and IQ was lower (p<0.001) in the FEP group compared to the HC group (Table 1). Use of cannabis and benzodiazepine was higher in the FEP group (p≤0.01), while alcohol use was more evenly distributed compared to the HCs (p<0.001). Patients gained more weight than HCs during the initial six weeks (p=0.01). Between the two treatment groups, the Amisulpride cohort had a higher increase in weight (p<0.001) and less reduction in negative symptoms (ΔPANSS_negative_, p=0.01) than the Aripiprazole cohort during the six-week treatment period. Use of cannabis was higher in the Amisulpride cohort (p=0.004).

**Figure 1:**
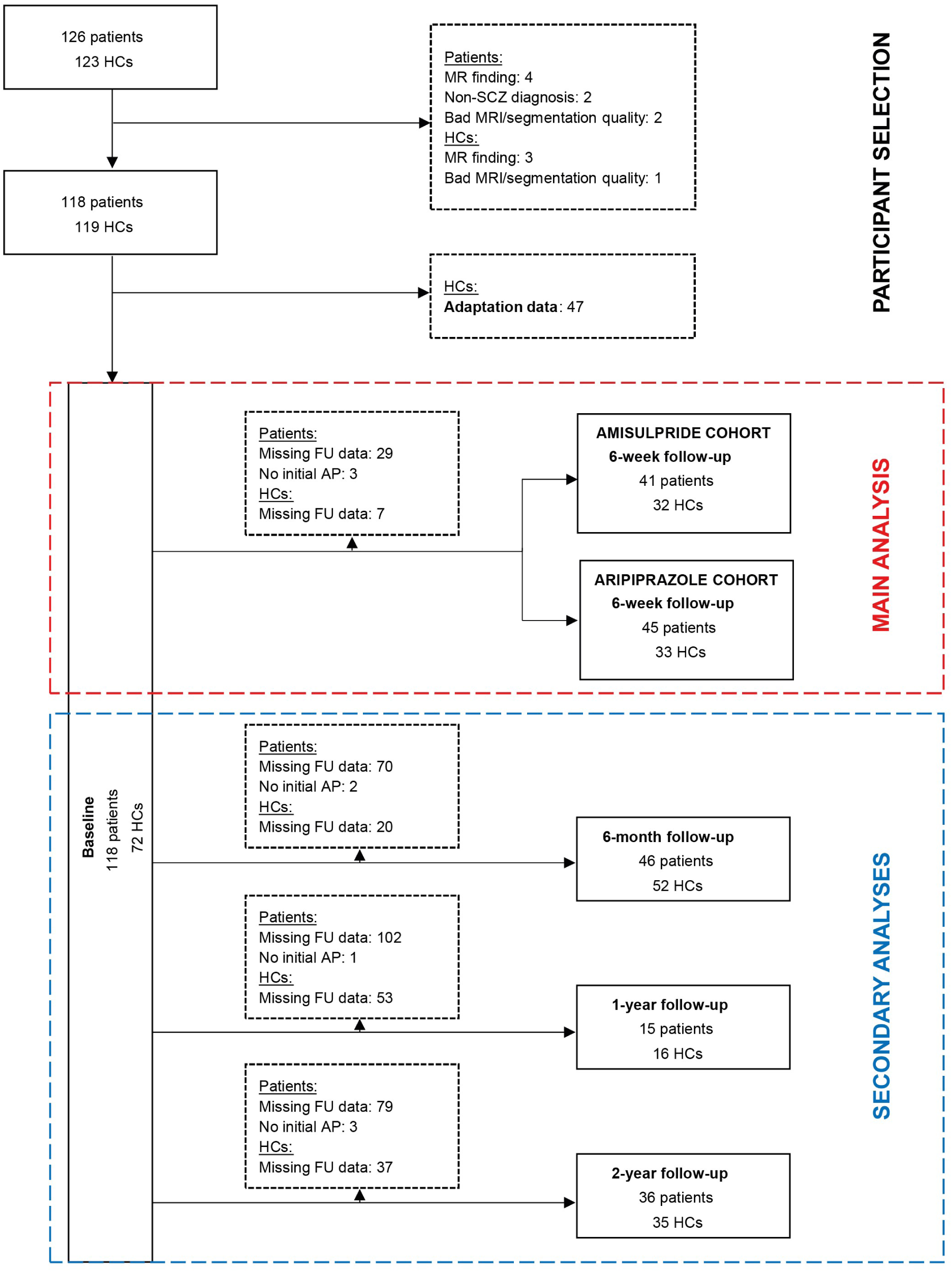
Flow chart of participant selection prior to model application and overview of participants included in main and secondary analyses. FU, follow-up.

**Table 1:**
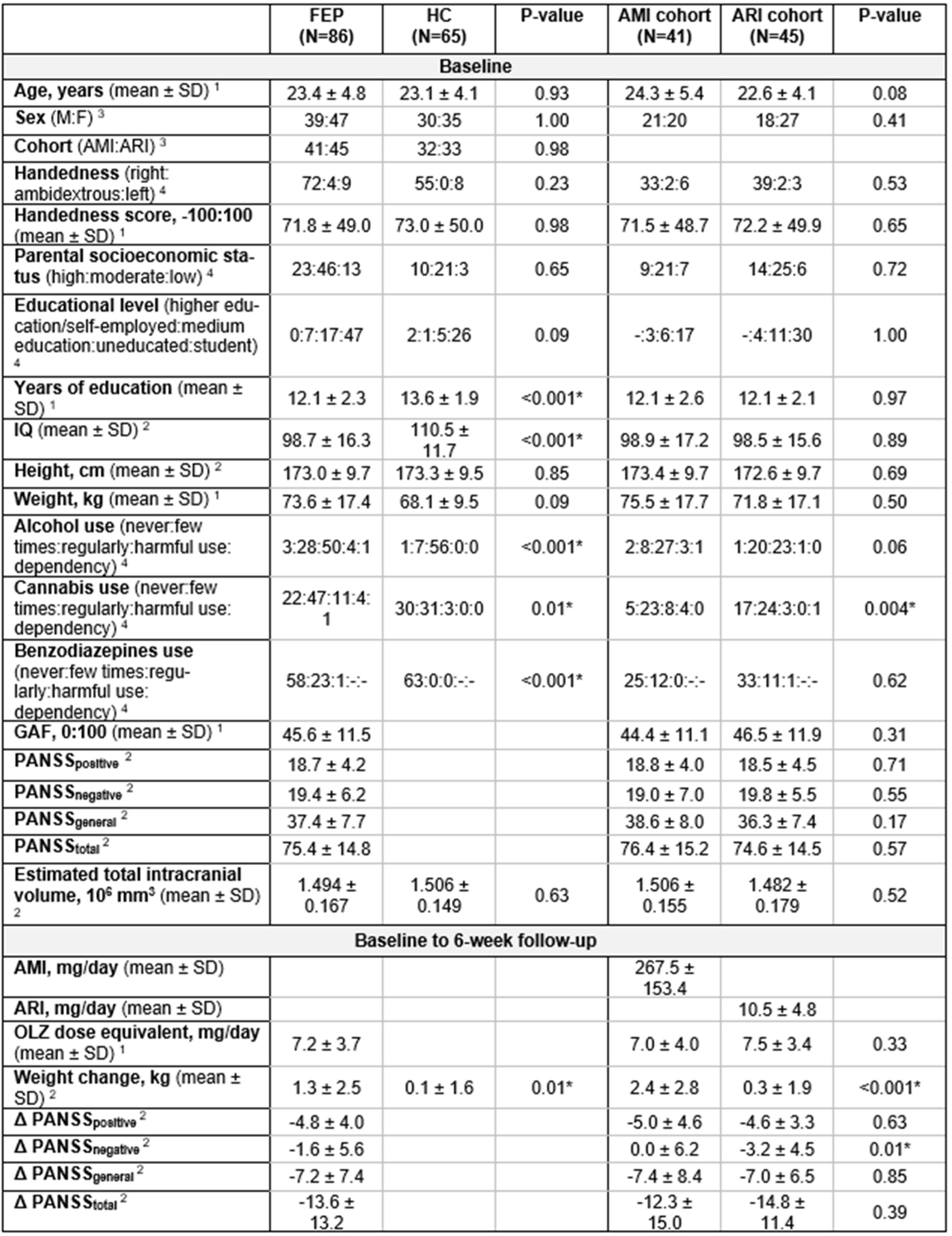
Demographic and clinical characteristics of participants included in the main analysis. AMI, amisulpride. ARI, aripiprazole. SD, standard deviation. GAF, Global Assessment of Functioning. OLZ, olanzapine. ^1^Mann-Whitney U-test, ^2^two-sample t-test, ^3^chi-squared test, ^4^Fisher’s exact test. Significant p-values are marked with *.

Patients who dropped out between baseline and the six-week follow-up visit had lower GAF scores (p<0.001), and higher PANSS subscale and total PANSS scores (p<0.04) at baseline compared to patients who continued to participate in the study (Supplementary Table S1).

In the secondary analyses of long-term changes (i.e., six months, one year, and two years), we included a total of 114 participants (101 of whom were also represented in the main analysis) with MRI data from baseline and at least one long-term follow-up. Demographic and clinical comparisons of groups of participants included in the secondary analyses are shown in Supplementary Table S2.

### Structural brain differences at baseline

At baseline, patients with six-week follow-up data (N=86) differed from HCs (N=65) in 13 IDPs (p_uncorrected_<0.05) (Supplementary Figure S1, Supplementary Table S3), but none remained significant after correction for multiple comparisons. Similarly, comparison of all patients (N=118) and HCs (N=72) with baseline data did not reveal any significant IDPs after FDR-correction.

### Structural changes after six weeks

#### Amisulpride cohort

Patients treated with amisulpride (N=41) displayed significant trajectory deviations in three IDPs after six weeks of monotherapy (Figure 2, Supplementary Table S4). Patients displayed striatal volume increases bilaterally in the caudate nucleus (left: 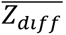=0.76, p=0.01; right: 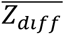=0.99, p=0.01) and in the right putamen (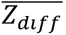=0.66, p=0.01). Patients displayed no cortical changes.

**Figure 2:**
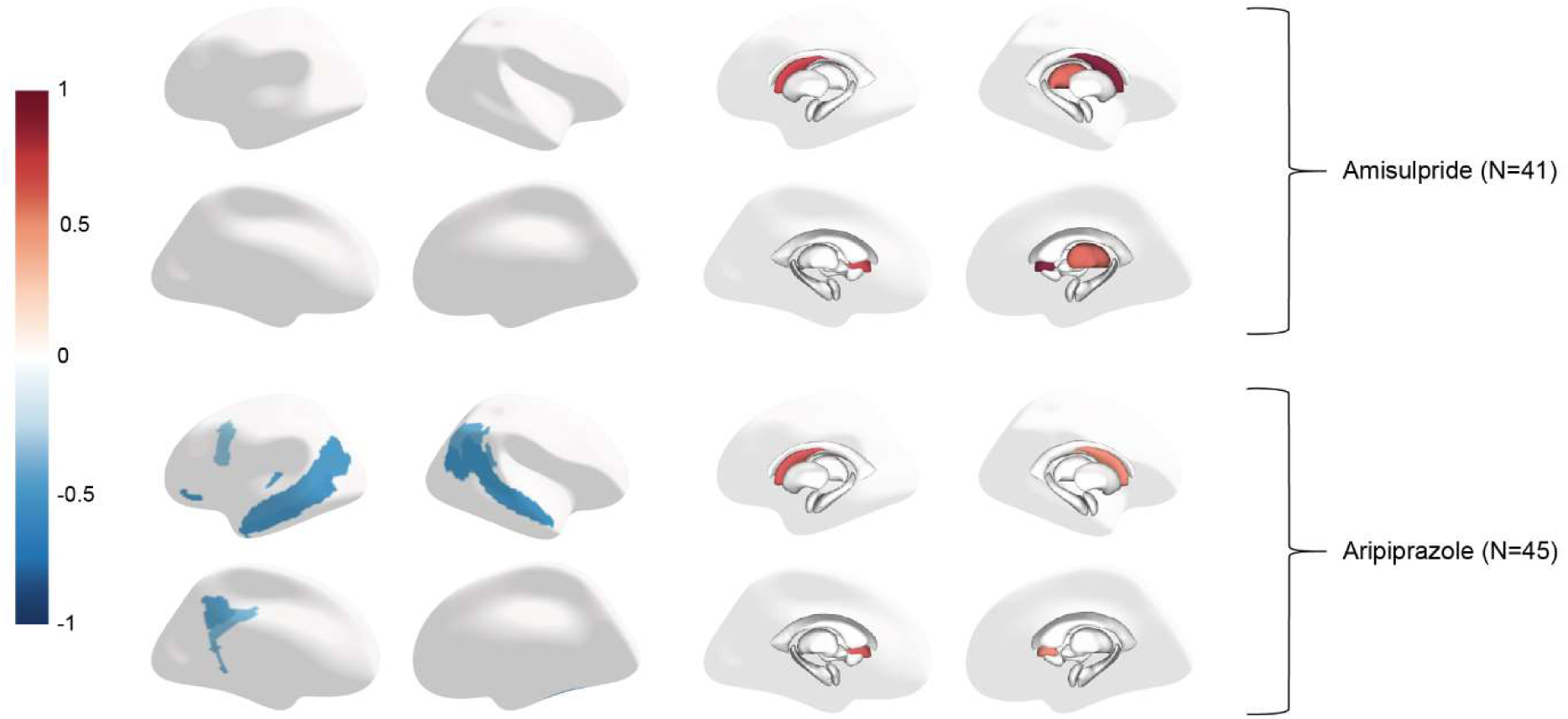
Cortical thickness and subcortical volume IDPs that deviate significantly (p<0.05) from their predicted trajectories in patients from baseline to six-week follow-up. The colormap reflects 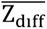.

#### Aripiprazole cohort

Patients treated with aripiprazole (N=45) displayed significant trajectory deviations in 13 IDPs after six weeks of monotherapy (Figure 2, Supplementary Table S5). Patients displayed volume increases in the bilateral caudate nucleus (left: 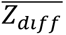=0.71, p <0.001; right: 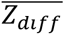=0.54, p=0.03). Patients also displayed cortical thinning in 11 regions, primarily in the bilateral temporal lobes, with 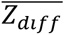 scores ranging from −0.60 to −0.40, p<0.05.

### Long-term structural changes after naturalistic treatment

#### From baseline to 6-month follow-up

After 6 months, patients (N=46) displayed significant trajectory deviations in 13 IDPs (Figure 3, Supplementary Table S6). Patients displayed increased volumes of the left caudate nucleus (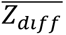=0.08, p=0.02) and right pallidum (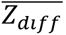=0.18, p=0.02). Patients displayed cortical thinning in 11 regions (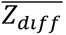 ranging from −0.71 to −0.12, p < 0.05).

**Figure 3:**
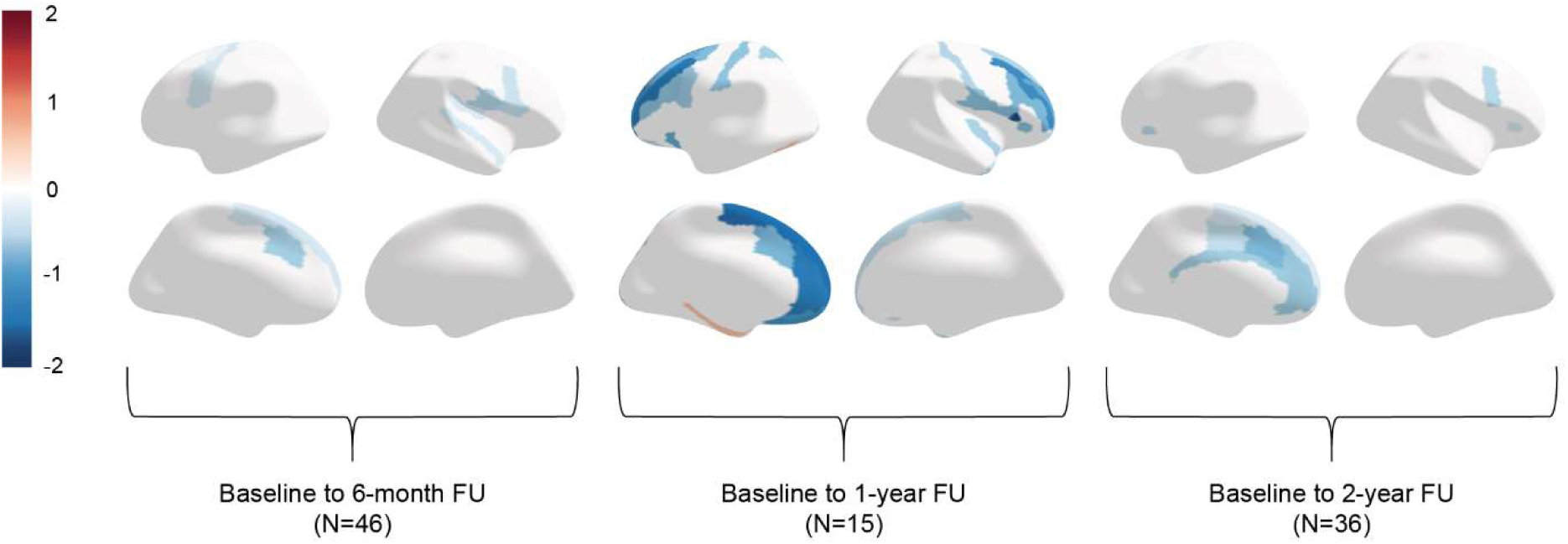
Cortical thickness IDPs that deviate significantly (p<0.05) from their predicted trajectories in patients from baseline to 6 months, 1 year, and 2 years, respectively. The colormap reflects 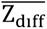. Patients only displayed subcortical volume changes after 6 months (not shown).

#### From baseline to 1-year follow-up

After 1 year, patients (N=15) displayed trajectory deviations in 40 IDPs (Figure 3, Supplementary Table S7). Patients displayed no subcortical changes, but increased volumes of the left cerebellum WM (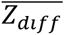=1.11, p=0.03) and the left choroid plexus (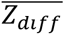=0.84, p=0.0477). Patients displayed cortical thinning in 35 regions (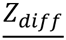 ranging from −1.88 to - 0.48, p<0.05), primarily located in the frontal lobes. Furthermore, increases in cortical thickness were identified in three regions in the left hemisphere with 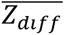 between 0.65 and 0.66, p<0.05.

#### From baseline to 2-year follow-up

After 2 years, the patients (N=36) displayed trajectory deviations in 11 IDPs (Figure 3, Supplementary Table S8). Patients displayed no subcortical volume changes but increased volume of the bilateral cerebellar WM (left: 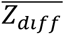=0.44, p=0.003; right: 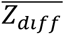 =0.46, p=0.001). Cortical thinning was found in nine regions (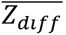 ranging from −0.15 to −0.95, p<0.05), mainly confined to the left medial frontal lobe.

#### Exploration of cohort biases after six weeks

In the Amisulpride cohort, a negative correlation was found between change in body weight and Z_diff_ scores in the caudate nucleus (ρ=-0.35, p_uncorrected_=0.03). In the Aripiprazole cohort, no correlations were found between change in body weight and Z_diff_ scores in any of the significant IDPs.

AP dose was not correlated with Z_diff_ scores in any significant IDPs in either cohort.

#### Correlations with symptom severity

The univariate approach yielded a positive correlation between Z_diff_ scores in the left ACC and ΔPANSS_general_ after 2 years (ρ=0.52, p<0.05) (Supplementary Figure S2). The pattern-based approach identified a principal component of change, characterized by negative weights in temporal and frontal regions (particularly the left ACC), which was negatively correlated with ΔPANSS_general_ after 2 years (ρ=-0.52, p<0.05) (Supplementary Figure S3). No other significant correlations between Z_diff_ scores and psychopathology emerged (see Supplementary Material for details).

## Discussion

In this study, we used a normative modeling approach to compare brain structural effects of amisulpride and aripiprazole in initially AP-naïve patients with FEP and to identify long-term changes after years of naturalistic treatment.

### Baseline structural deviations

At baseline, patients did not differ from controls in any cortical IDP, which is consistent with previous studies conducted on the same cohorts (46,47). However, regional thinner cortex and smaller hippocampal volumes were observed at trend-level (p_uncorrected_<0.05<p). In line with this observation, other studies have reported lower hippocampal volumes in AP-naïve FEP patients (48), and either widespread cortical thinning (49–51) or no differences from controls (52). Discrepancies may be attributed to varying patient samples and methods.

### Short-term structural effects of antipsychotics

After six weeks of AP monotherapy, both patient cohorts displayed striatal volume increases. In the Amisulpride cohort, the affected striatal regions were the bilateral caudate nucleus and the right putamen. Changes in these regions, and absence of cortical changes, are consistent with previous results obtained using conventional methods (17,46). In the Aripiprazole cohort, patients showed volumetric increases in the bilateral caudate nucleus and cortical thinning in the temporal lobe. Administration of comparable doses of amisulpride and aripiprazole for one week has recently been shown to cause similar striatal volume increases in healthy volunteers (53). To the best of our knowledge, no previous studies have investigated the structural effects of aripiprazole in AP-naïve FEP patients. Interestingly, a negative correlation was found between volume change in the left caudate nucleus and change in body weight in the Amisulpride cohort. To our knowledge, AP-associated changes in caudate nucleus volume have not previously been tied to body weight, and future research is needed to address the reliability of this association.

When comparing the two cohorts, the Amisulpride cohort exhibited numerically more striatal regions with significant changes than the Aripiprazole cohort. This is corroborated by a study on healthy people, where amisulpride induced increases in the right caudate nucleus and left putamen, while aripiprazole only induced increases in the right putamen (53). Collectively, these results suggest that D2R antagonism may cause more extensive striatal volume increases than partial D2R agonism. Conversely, the observed cortical thinning in the temporal lobe does not appear to result from functional D2R antagonism but may rather result from either functional D2R agonism or binding to one of the other targets of aripiprazole (54). For instance, aripiprazole targets serotonergic receptors through either antagonism (5-HT_2A_) or partial agonism (5-HT_1A_) (54). In patients with major depressive disorder, eight weeks of treatment with selective serotonin reuptake inhibitors (SSRIs), which exert their effect by increasing (synaptic) serotonin activity, has been shown to increase cortical thickness (55,56). Therefore, aripiprazole-mediated blockade of serotonergic receptors, counteracting the effects of serotonin, could be the cause of cortical thinning. Finally, the previously mentioned increases in resting striatal blood flow in the Aripiprazole cohort (20,21) appear to coincide with striatal volumetric increases. For this reason, it could also be speculated whether the two are causally related.

### Long-term structural changes

After six months, patients displayed volume increases in the left caudate nucleus and right pallidum. The latter has also been reported enlarged in initially AP-naïve FEP patients after three months of SGA treatment (16). Interestingly, no striatal volume increases were found at later follow-ups (1 and 2 years), thus reflecting a return to their baseline trajectories. Interestingly, a similar pattern has been observed for striatal perfusion with striatal increases after 6 weeks and 6 months but a normalization after two years (21). This return could be the result of biological compensatory mechanisms, such as changes in dopamine receptor expressions. In support of this, repeated aripiprazole administration has been found to cause D2R upregulation in rats (57). In schizophrenia patients, years of AP exposure has also been associated with increased D2R binding potential (58).

Our patients displayed widespread cortical thinning after 6 months. After one year, cortical thinning was mainly observed in the bilateral frontal lobes, and after 2 years, it was confined to the left medial frontal lobe and cingulate cortex. The predominantly observed thinning following naturalistic treatment is in accordance with a large body of literature reporting progressive frontal thinning in patients treated with first or second generation APs (59,60). However, the magnitude of thickness reductions observed at our 1- and 2-year follow-ups contradict the proposed progressive manner of thinning. Newer studies employing normative modeling approaches on data from FEP patients report attenuation of baseline cortical thinning years after inclusion (30,61). In partial compliance with this, we also identified limited increases in cortical thickness after 1 year. The lack of prior AP treatment at baseline distinguishes our sample from those previously studied, and perhaps this constitutes an explanation for the inconsistencies with existing literature. Generally, subtle differences in patient samples combined with high heterogeneity complicates inter-study comparisons. Due to observational cohort study design the absence of an untreated patient group for comparison limits inferences regarding causality between antipsychotic exposure and brain structural effects. This limitation is particularly relevant when disentangling long-term brain changes from illness-related factors. In accordance, previous data has linked long-term cortical thinning to combined effects of AP treatment and illness progression (62).

We found that after two years, ΔPANSS_general_ was correlated with both Z_diff_ scores in the left ACC and a principal component of change, which was strongly influenced by the left ACC. Accordingly, the ACC has been linked to cognitive and emotional regulation (63) and its activation associated with successful processing of emotional stimuli (64). Similarly, less cortical thinning could in this case reflect better neurofunctional management of the psychosocial stressors associated with longer illness duration. While our correlation results have limited impact, future research should address the external validity and potential confounding effects of cumulative AP dose.

### Strengths and limitations

This study has several major strengths, including the longitudinal study design with multiple follow-ups with a large group of initially AP-naïve patients. This allowed us to rule out structural correlates of prior AP treatment and, combined with a normative modeling approach, enabled quantification of within-subject changes relative to the predicted structural trajectories. The use of monotherapy with either amisulpride or aripiprazole further permitted investigation of their shared and distinct structural effects. Although the sample size was moderate at baseline, it was reduced throughout the multiple follow-ups. Therefore, the sample size and varying availability of follow-up data is a limitation to this study. We found that participants who dropped out of the study before the six-week follow-up visit had more severe symptoms and were more functionally impaired at baseline. Thus, the dropout was selective, and this might have implications for the generalizability of our results.

## Conclusions

In this study, we confirmed transient striatal volume increases in initially AP-naïve FEP patients treated with amisulpride and identified similar increases in patients treated with aripiprazole. In addition, short-term aripiprazole monotherapy was found to induce temporal thinning. This study shows the importance of considering the pharmacological profile of APs when investigating their effects on brain structure. Secondarily, we identified cortical thinning in the frontal lobes of patients after years of naturalistic treatment, likely reflecting the combined structural effects of pharmacological treatment and illness progression in patients with psychosis.

## Author contributions

BE, KA, and AF conceived the study. BG conceived the original cohort studies, and KB and MN collected the data. JR contributed with MRI data processing. BB and AM developed the normative model. AF conducted the statistical analyses. AF and KS drafted the manuscript. KA, WS, JR, and BB particularly contributed with method section. BG, BE, KB, and AM particularly contributed with interpretation and discussion. All authors fulfill the authorship criteria of the IMCJE and have provided feedback and approval of the final manuscript.

## Data availability

The datasets analyzed during the current study are not publicly available to protect the privacy of research participants but are available from the corresponding author on reasonable request. Statistical code is available upon request.

## References

1. McCutcheon RA, Reis Marques T, Howes OD. Schizophrenia—An Overview. JAMA Psychiatry. 1. februar 2020;77(2):201.

2. Wannan CMJ, Cropley VL, Chakravarty MM, Bousman C, Ganella EP, Bruggemann JM, m.fl. Evidence for Network-Based Cortical Thickness Reductions in Schizophrenia. Am J Psychiatry. 1. juli 2019;176(7):552–63.

3. Zhao Y, Zhang Q, Shah C, Li Q, Sweeney JA, Li F, m.fl. Cortical Thickness Abnormalities at Different Stages of the Illness Course in Schizophrenia: A Systematic Review and Meta-analysis. JAMA Psychiatry. 1. juni 2022;79(6):560.

4. Fan F, Xiang H, Tan S, Yang F, Fan H, Guo H, m.fl. Subcortical structures and cognitive dysfunction in first episode schizophrenia. Psychiatry Res Neuroimaging. april 2019;286:69–75.

5. Fang K, Wen B, Niu L, Wan B, Zhang W. Higher brain structural heterogeneity in schizophrenia. Front Psychiatry. 23. september 2022;13:1017399.

6. Shenton ME, Dickey CC, Frumin M, McCarley RW. A review of MRI findings in schizophrenia. Schizophr Res 2001;49(1–2):1–52.

7. Howes OD, Kapur S. The Dopamine Hypothesis of Schizophrenia: Version III--The Final Common Pathway. Schizophr Bull. 30. marts 2009;35(3):549–62.

8. Kaar SJ, Natesan S, McCutcheon R, Howes OD. Antipsychotics: Mechanisms underlying clinical response and side-effects and novel treatment approaches based on pathophysiology. Neuropharmacology. august 2020;172:107704.

9. Van Erp TGM, Walton E, Hibar DP, Schmaal L, Jiang W, Glahn DC, m.fl. Cortical Brain Abnormalities in 4474 Individuals With Schizophrenia and 5098 Control Subjects via the Enhancing Neuro Imaging Genetics Through Meta Analysis (ENIGMA) Consortium. Biol Psychiatry. november 2018;84(9):644–54.

10. Haijma SV, Van Haren N, Cahn W, Koolschijn PCMP, Hulshoff Pol HE, Kahn RS. Brain Volumes in Schizophrenia: A Meta-Analysis in Over 18 000 Subjects. Schizophr Bull. september 2013;39(5):1129–38.

11. Veijola J, Guo JY, Moilanen JS, Jääskeläinen E, Miettunen J, Kyllönen M, m.fl. Longitudinal Changes in Total Brain Volume in Schizophrenia: Relation to Symptom Severity, Cognition and Antipsychotic Medication. Takei N, redaktør. PLoS ONE. 18. juli 2014;9(7):e101689.

12. Chakos MH, Shirakawa O, Lieberman J, Lee H, Bilder R, Tamminga CA. Striatal enlargement in rats chronically treated with neuroleptic. Biol Psychiatry. oktober 1998;44(8):675–84.

13. Andersson C. Striatal Volume Changes in the Rat Following Long-term Administration of Typical and Atypical Antipsychotic Drugs. Neuropsychopharmacology. august 2002;27(2):143–51.

14. Vernon AC, Natesan S, Crum WR, Cooper JD, Modo M, Williams SCR, m.fl. Contrasting Effects of Haloperidol and Lithium on Rodent Brain Structure: A Magnetic Resonance Imaging Study with Postmortem Confirmation. Biol Psychiatry. maj 2012;71(10):855–63.

15. Glenthoj A, Glenthoj BY, Mackeprang T, Pagsberg AK, Hemmingsen RP, Jernigan TL, m.fl. Basal ganglia volumes in drug-naive first-episode schizophrenia patients before and after short-term treatment with either a typical or an atypical antipsychotic drug. Psychiatry Res Neuroimaging. april 2007;154(3):199–208.

16. Chopra S, Fornito A, Francey SM, O’Donoghue B, Cropley V, Nelson B, m.fl. Differentiating the effect of antipsychotic medication and illness on brain volume reductions in first-episode psychosis: A Longitudinal, Randomised, Triple-blind, Placebo-controlled MRI Study. Neuropsychopharmacology. juli 2021;46(8):1494–501.

17. Andersen HG, Raghava JM, Svarer C, Wulff S, Johansen LB, Antonsen PK, m.fl. Striatal Volume Increase After Six Weeks of Selective Dopamine D2/3 Receptor Blockade in First-Episode, Antipsychotic-Naïve Schizophrenia Patients. Front Neurosci. 20. maj 2020;14:484.

18. Orzelska-Górka J, Mikulska J, Wiszniewska A, Biała G. New Atypical Antipsychotics in the Treatment of Schizophrenia and Depression. Int J Mol Sci. 13. september 2022;23(18):10624.

19. Medicinrådet. Medicinrådets behandlingsvejledning vedrørende antipsykotika til behandling af psykotiske tilstande hos voksne [Internet]. Medicinrådet; 2020 jul. Tilgængelig hos: https://medicinraadet.dk/anbefalinger-og-vejledninger/behandlingsvejledninger-og-laegemiddelrekommandationer/antipsykotika-til-voksne

20. Bojesen KB, Glenthøj BY, Sigvard AK, Tangmose K, Raghava JM, Ebdrup BH, m.fl. Cerebral blood flow in striatum is increased by partial dopamine agonism in initially antipsychotic-naïve patients with psychosis. Psychol Med. oktober 2023;53(14):6691–701.

21. Bojesen KB, Rostrup E, Sigvard AK, Mikkelsen M, Edden RAE, Ebdrup BH, m.fl. The Trajectory of Prefrontal GABA Levels in Initially Antipsychotic-Naïve Patients With Psychosis During 2 Years of Treatment and Associations With Striatal Cerebral Blood Flow and Outcome. Biol Psychiatry Cogn Neurosci Neuroimaging. juli 2024;9(7):703–13.

22. Ho BC, Andreasen NC, Ziebell S, Pierson R, Magnotta V. Long-term Antipsychotic Treatment and Brain Volumes: A Longitudinal Study of First-Episode Schizophrenia. Arch Gen Psychiatry. 7. februar 2011;68(2):128.

23. Marquand AF, Rezek I, Buitelaar J, Beckmann CF. Understanding Heterogeneity in Clinical Cohorts Using Normative Models: Beyond Case-Control Studies. Biol Psychiatry. oktober 2016;80(7):552–61.

24. Marquand AF, Kia SM, Zabihi M, Wolfers T, Buitelaar JK, Beckmann CF. Conceptualizing mental disorders as deviations from normative functioning. Mol Psychiatry. oktober 2019;24(10):1415–24.

25. Zabihi M, Oldehinkel M, Wolfers T, Frouin V, Goyard D, Loth E, m.fl. Dissecting the Heterogeneous Cortical Anatomy of Autism Spectrum Disorder Using Normative Models. Biol Psychiatry Cogn Neurosci Neuroimaging. juni 2019;4(6):567–78.

26. Wolfers T, Beckmann CF, Hoogman M, Buitelaar JK, Franke B, Marquand AF. Individual differences v. the average patient: mapping the heterogeneity in ADHD using normative models. Psychol Med. januar 2020;50(2):314–23.

27. Zabihi M, Floris DL, Kia SM, Wolfers T, Tillmann J, Arenas AL, m.fl. Fractionating autism based on neuroanatomical normative modeling. Transl Psychiatry. 6. november 2020;10(1):384.

28. Wolfers T, Rokicki J, Alnæs D, Berthet P, Agartz I, Kia SM, m.fl. Replicating extensive brain structural heterogeneity in individuals with schizophrenia and bipolar disorder. Hum Brain Mapp. juni 2021;42(8):2546–55.

29. Lv J, Di Biase M, Cash RFH, Cocchi L, Cropley VL, Klauser P, m.fl. Individual deviations from normative models of brain structure in a large cross-sectional schizophrenia cohort. Mol Psychiatry. juli 2021;26(7):3512–23.

30. Bučková BR, Fraza C, Rehák R, Kolenič M, Beckmann CF, Španiel F, m.fl. Using normative models pre-trained on cross-sectional data to evaluate intra-individual longitudinal changes in neuroimaging data. eLife. 12. marts 2025;13:RP95823.

31. Rutherford S, Fraza C, Dinga R, Kia SM, Wolfers T, Zabihi M, m.fl. Charting brain growth and aging at high spatial precision. eLife. 1. februar 2022;11:e72904.

32. Nielsen MØ, Rostrup E, Wulff S, Bak N, Lublin H, Kapur S, m.fl. Alterations of the Brain Reward System in Antipsychotic Naïve Schizophrenia Patients. Biol Psychiatry. maj 2012;71(10):898–905.

33. Bojesen KB, Ebdrup BH, Jessen K, Sigvard A, Tangmose K, Edden RAE, m.fl. Treatment response after 6 and 26 weeks is related to baseline glutamate and GABA levels in antipsychotic-naïve patients with psychosis. Psychol Med. oktober 2020;50(13):2182–93.

34. Leucht S, Samara M, Heres S, Patel MX, Furukawa T, Cipriani A, m.fl. Dose Equivalents for Second-Generation Antipsychotic Drugs: The Classical Mean Dose Method. Schizophr Bull. november 2015;41(6):1397–402.

35. Kay SR, Fiszbein A, Opler LA. The Positive and Negative Syndrome Scale (PANSS) for Schizophrenia. Schizophr Bull. 1. januar 1987;13(2):261–76.

36. Wechsler D. Manual for the Wechsler Adult Intelligence Scale--Third Edition (WAIS-III). San Antonio: The Psychological Corporation; 1997.

37. Oldfield RC. The assessment and analysis of handedness: The Edinburgh inventory. Neuropsychologia. marts 1971;9(1):97–113.

38. Dale AM, Fischl B, Sereno MI. Cortical Surface-Based Analysis: I. Segmentation and surface reconstruction. NeuroImage. februar 1999;9(2):179–94.

39. Fischl B, Sereno MI, Dale AM. Cortical Surface-Based Analysis: II. Inflation, flattening, and a surface-based coordinate system. NeuroImage. februar 1999;9(2):195–207.

40. Reuter M, Fischl B. Avoiding asymmetry-induced bias in longitudinal image processing. NeuroImage. juli 2011;57(1):19–21.

41. Destrieux C, Fischl B, Dale A, Halgren E. Automatic parcellation of human cortical gyri and sulci using standard anatomical nomenclature. NeuroImage. oktober 2010;53(1):1–15.

42. Fischl B, Salat DH, Busa E, Albert M, Dieterich M, Haselgrove C, m.fl. Whole Brain Segmentation: automated labeling of neuroanatomical structures in the human brain. Neuron. januar 2002;33(3):341–55.

43. Klapwijk ET, Van De Kamp F, Van Der Meulen M, Peters S, Wierenga LM. Qoala-T: A supervised-learning tool for quality control of FreeSurfer segmented MRI data. NeuroImage. april 2019;189:116–29.

44. Fraza CJ, Dinga R, Beckmann CF, Marquand AF. Warped Bayesian linear regression for normative modelling of big data. NeuroImage. december 2021;245:118715.

45. Benjamini Y, Hochberg Y. Controlling the False Discovery Rate: A Practical and Powerful Approach to Multiple Testing. J R Stat Soc Ser B Stat Methodol. 1. januar 1995;57(1):289–300.

46. Jessen K, Rostrup E, Mandl RCW, Nielsen MØ, Bak N, Fagerlund B, m.fl. Cortical structures and their clinical correlates in antipsychotic-naïve schizophrenia patients before and after 6 weeks of dopamine D_2/3_ receptor antagonist treatment. Psychol Med. april 2019;49(5):754–63.

47. Jessen K, Mandl RCW, Fagerlund B, Bojesen KB, Raghava JM, Obaid HG, m.fl. Patterns of Cortical Structures and Cognition in Antipsychotic-Naïve Patients With First-Episode Schizophrenia: A Partial Least Squares Correlation Analysis. Biol Psychiatry Cogn Neurosci Neuroimaging. maj 2019;4(5):444–53.

48. Ebdrup BH, Glenthøj B, Rasmussen H, Aggernaes B, Langkilde AR, Paulson OB, m.fl. Hippocampal and caudate volume reductions in antipsychotic-naive first-episode schizophrenia. J Psychiatry Neurosci. marts 2010;35(2):95–104.

49. Song X, Quan M, Lv L, Li X, Pang L, Kennedy D, m.fl. Decreased cortical thickness in drug naïve first episode schizophrenia: In relation to serum levels of BDNF. J Psychiatr Res. januar 2015;60:22–8.

50. Xiao Y, Lui S, Deng W, Yao L, Zhang W, Li S, m.fl. Altered Cortical Thickness Related to Clinical Severity But Not the Untreated Disease Duration in Schizophrenia. Schizophr Bull. 1. januar 2015;41(1):201–10.

51. Plitman E, Patel R, Chung JK, Pipitone J, Chavez S, Reyes-Madrigal F, m.fl. Glutamatergic Metabolites, Volume and Cortical Thickness in Antipsychotic-Naive Patients with First-Episode Psychosis: Implications for Excitotoxicity. Neuropsychopharmacology. september 2016;41(10):2606–13.

52. Rais M, Cahn W, Schnack HG, Hulshoff Pol HE, Kahn RS, Van Haren NEM. Brain volume reductions in medication-naive patients with schizophrenia in relation to intelligence quotient. Psychol Med. september 2012;42(9):1847–56.

53. Selvaggi P, Osugo M, Zahid U, Dipasquale O, Whitehurst T, Onwordi E, m.fl. Antipsychotics cause reversible structural brain changes within one week. Neuropsychopharmacology [Internet]. 7. maj 2025 [henvist 8. maj 2025]; Tilgængelig hos: https://www.nature.com/articles/s41386-025-02120-4

54. Shapiro DA, Renock S, Arrington E, Chiodo LA, Liu LX, Sibley DR, m.fl. Aripiprazole, A Novel Atypical Antipsychotic Drug with a Unique and Robust Pharmacology. Neuropsychopharmacology. august 2003;28(8):1400–11.

55. Koenig J, Westlund Schreiner M, Klimes-Dougan B, Ubani B, Mueller BA, Lim KO, m.fl. Increases in orbitofrontal cortex thickness following antidepressant treatment are associated with changes in resting state autonomic function in adolescents with major depression – Preliminary findings from a pilot study. Psychiatry Res Neuroimaging. november 2018;281:35–42.

56. Nemati S, Abdallah CG. Increased Cortical Thickness in Patients With Major Depressive Disorder Following Antidepressant Treatment. Chronic Stress. januar 2020;4:2470547019899962.

57. Varela FA, Der-Ghazarian T, Lee RJ, Charntikov S, Crawford CA, McDougall SA. Repeated aripiprazole treatment causes dopamine D2 receptor up-regulation and dopamine supersensitivity in young rats. J Psychopharmacol (Oxf). april 2014;28(4):376–86.

58. Silvestri S, Seeman MV, Negrete JC, Houle S, Shammi CM, Remington GJ, m.fl. Increased dopamine D 2 receptor binding after long-term treatment with antipsychotics in humans: a clinical PET study. Psychopharmacology (Berl). 6. oktober 2000;152(2):174–80.

59. Gutiérrez-Galve L, Chu EM, Leeson VC, Price G, Barnes TRE, Joyce EM, m.fl. A longitudinal study of cortical changes and their cognitive correlates in patients followed up after first-episode psychosis. Psychol Med. januar 2015;45(1):205–16.

60. Van Haren NEM. Changes in Cortical Thickness During the Course of Illness in Schizophrenia. Arch Gen Psychiatry. 1. september 2011;68(9):871.

61. Berthet P, Haatveit BC, Kjelkenes R, Worker A, Kia SM, Wolfers T, m.fl. A 10-Year Longitudinal Study of Brain Cortical Thickness in People with First-Episode Psychosis Using Normative Models. Schizophr Bull. 20. december 2024;51(1):95–107.

62. Fusar-Poli P, Smieskova R, Kempton MJ, Ho BC, Andreasen NC, Borgwardt S. Progressive brain changes in schizophrenia related to antipsychotic treatment? A meta-analysis of longitudinal MRI studies. Neurosci Biobehav Rev. september 2013;37(8):1680–91.

63. Bush G, Luu P, Posner MI. Cognitive and emotional influences in anterior cingulate cortex. Trends Cogn Sci. juni 2000;4(6):215–22.

64. Shin LM, Whalen PJ, Pitman RK, Bush G, Macklin ML, Lasko NB, m.fl. An fMRI study of anterior cingulate function in posttraumatic stress disorder. Biol Psychiatry. december 2001;50(12):932–42.

