## Supplementary Material for "Comparing brain structural effects of dopaminergic antagonism and partial agonism in antipsychotic-naïve patients with first-episode psychosis using normative modeling"

### Details on deviation (Z and $Z_{diff}$) scores

For the *n*'th subject and *d*'th IDP, the Z-score is calculated by

$$Z_{nd} = \frac{y_{nd} - \widehat{y_{nd}}}{\sqrt{{\sigma_d}^2 + ({\sigma_*}^2)_d}},$$

where $y_{nd}$ is the true response variable, $\widehat{y_{nd}}$ is the predictive mean, and the denominator is the standard deviation based on the estimated noise variance, ${\sigma_d}^2$, and the variance caused by modeling uncertainty for the *d*'th IDP, $({\sigma_*}^2)_d$ (1,2)

For a subject *n* in IDP *d*, the $Z_{diff}$ score can be computed by

$$Z_{diff,nd} = \frac{\Delta y_{nd} - \widehat{\Delta y_{nd}}}{\sqrt{{\sigma_v}^2 + ({\sigma_{\Delta *}}^2)_d}},$$

where $\Delta y_{nd}$ is the change in the true response variable, $\widehat{\Delta y_{nd}}$ is the change in the predictive mean, and the denominator is the standard deviation, consisting of the visit-specific noise variance $({\sigma_v}^2)$ and the variance attributed to modeling uncertainty $(({\sigma_{\Delta *}}^2)_d)$ (3). The denominator was estimated by computing the variance of the numerator for the remaining (i.e., non-adaptation) HCs.

### Correlation analyses

We explored the correlations between structural changes and both treatment response and potential cohort biases using Spearman's rank-order correlation. For all longitudinal analyses, the correlation with treatment response was investigated using two different approaches. Firstly, using a univariate approach, we calculated the correlations between $Z_{diff}$ scores in significant IDPs and the accompanying changes in PANSS subscales ($\Delta PANSS_{positive}$, $\Delta PANSS_{negative}$, and $\Delta PANSS_{general}$). Secondly, using a pattern-based approach, we reduced the dimensionality of $Z_{diff}$ scores, i.e., across IDPs, using Principal Component Analysis (PCA) applied to data from both patients and HCs. Subsequently, we determined the correlations between each principal component and changes in PANSS subscales. Only principal components with an explained variance ratio (EVR) ≥ 0.05 were included in the correlation analyses. To address potential biases of the structural changes after six weeks, we explored correlations between $Z_{diff}$ scores and change in body weight and AP dose within each cohort.

#### *Correlations with symptom severity*

##### *Univariate approach*

The univariate approach yielded positive correlations ($p_{uncorrected}<0.05$) between significantly changed IDPs and changes in PANSS subscales at 6 months, 1 year, and 2 years (Supplementary Figure S2). Only the correlation between $Z_{diff}$ scores in the left ACC and $\Delta PANSS_{general}$ after 2 years remained significant after correction for multiple comparisons ($p<0.05$).

##### *Pattern-based approach*

In the pattern-based approach, 3-5 PCs were selected for each of the five longitudinal analyses. This approach yielded a total of three correlations ($p_{uncorrected}<0.05$) between PCs and changes in PANSS subscales at 6 months (PC4, EVR=0.05), 1 year (PC3, EVR=0.08), and 2 years (PC3, EVR=0.09) (Supplementary Figure S3). Only the correlation between PC3 and $\Delta PANSS_{general}$ after 2 years remained significant after correction for multiple comparisons ($p<0.05$).

### Supplementary Figures

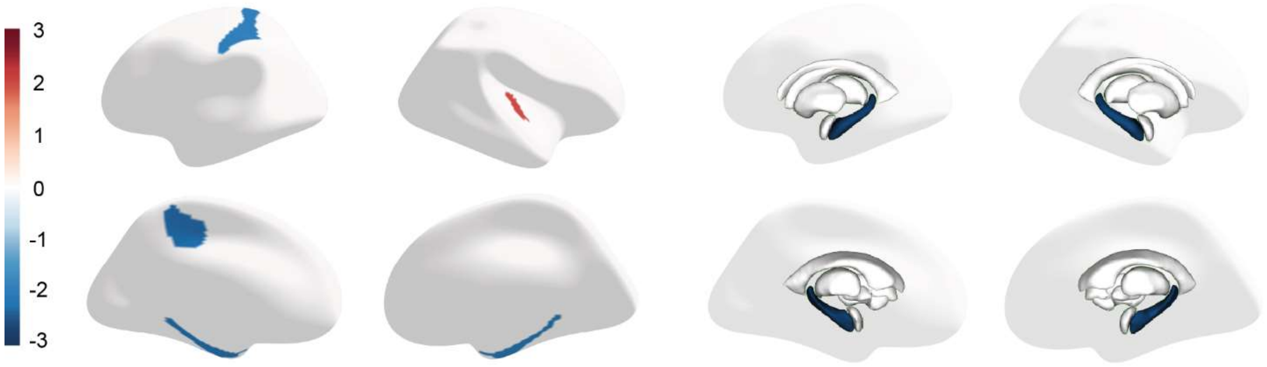


**Supplementary Figure S1:** Cortical thickness and subcortical volume IDPs, where the baseline Z-scores differ ($p<0.05$) between FEP patients (N=86) and HCs (N=65) at baseline. The colormap reflects the t-statistic. Negative values indicate that the mean Z-score is lower in the FEP group, while positive values indicate higher mean Z-score in the FEP group. None of the IDPs remained significant after correction for multiple testing.

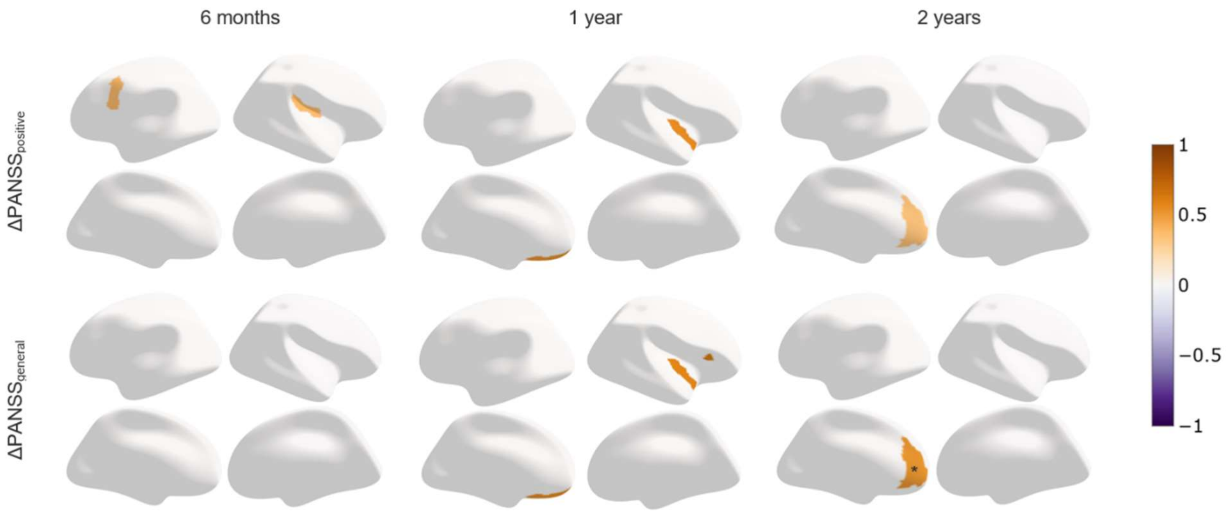


***Supplementary Figure S2**: Results of univariate approach. Only deviated IDPs where $Z_{diff}$ scores correlate ($p_{uncorrected}$<0.05) with changes in PANSS subscales are shown. The colormap reflects Spearman's correlation coefficient, ρ. Significant correlations after FDR-correction (p<0.05) are marked with *.*

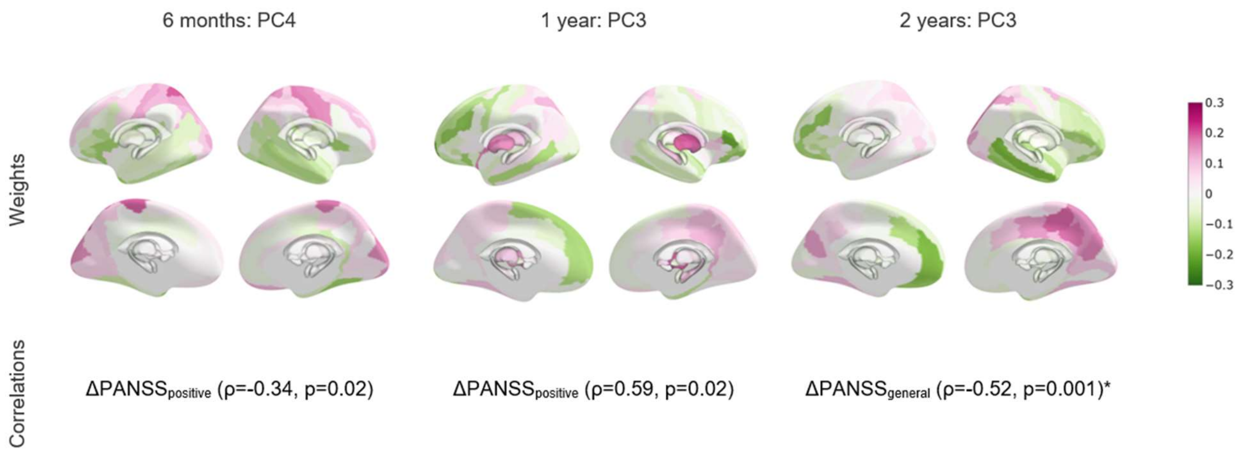


***Supplementary Figure S3**: Results of pattern-based approach. Only the PCs that correlate ($p_{uncorrected}$<0.05) with changes in PANSS subscales are shown. The colormap reflects the weight of each IDP in the PC. Significant correlations after FDR-correction (p<0.05) are marked with *.*

### Supplementary Tables

***Supplementary Table S1:*** *Demographic and clinical characteristics of patients with and without six-week follow-up data. AMI, amisulpride. ARI, aripiprazole. SD, standard deviation. GAF, Global Assessment of Functioning. OLZ, olanzapine. [1] Mann-Whitney U-test, [2] two-sample t-test, [3] chi-squared test, [4] Fisher's exact test. Significant p-values are marked with *.*

| | Patients with six-week FU (N=86) | Patients without six-week FU (N=32) | P-value |
|---|---|---|---|
| **Baseline** | | | |
| **Age, years** (mean ± SD) [1] | 23.4 ± 4.8 | 25.1 ± 6.4 | 0.32 |
| **Sex** (M:F) [3] | 39:47 | 21:11 | 0.08 |
| **Cohort** (AMI:ARI) [3] | 41:45 | 20:12 | 0.22 |
| **Handedness** (right: ambidextrous:left) [4] | 72:4:9 | 26:2:3 | 0.91 |
| **Handedness score, -100:100** (mean ± SD) [1] | 71.8 ± 49.0 | 67.7 ±51.5 | 0.37 |
| **Parental socioeconomic status** (high:moderate:low) [4] | 23:46:13 | 9:10:8 | 0.16 |
| **Educational level** (higher education/self-employed:medium education:uneducated:student) [4] | 0:7:17:47 | 3:4:8:13 | 0.03* |
| **Years of education** (mean ± SD)[1] | 12.1 ± 2.3 | 12.4 ± 2.5 | 0.46 |
| **IQ** (mean ± SD) [2] | 98.7 ± 16.3 | 100.0 ± 22.0 | 0.75 |
| **Height, cm** (mean ± SD) [2] | 173.0 ± 9.7 | 174.0 ± 10.2 | 0.63 |
| **Weight, kg** (mean ± SD) [1] | 73.6 ± 17.4 | 73.7 ± 16.4 | 0.93 |
| **Alcohol use** (never:few times:regularly:harmful use: dependency) [4] | 3:28:50:4:1 | 1:8:19:2:0 | 0.91 |
| **Cannabis use** (never:few times:regularly:harmful use: dependency) [4] | 22:47:11:4:1 | 7:14:7:3:0 | 0.50 |
| **Benzodiazepines use** (never:few times:regularly:harmful use: dependency) [4] | 58:23:1:-:- | 18:10:2:-:- | 0.18 |
| **GAF, 0:100** (mean ± SD) [1] | 45.6 ± 11.5 | 36.2 ± 9.0 | <0.001* |
| **$PANSS_{positive}$** (mean ± SD) [2] | 18.7 ± 4.2 | 20.6 ± 3.7 | 0.03 |
| **$PANSS_{negative}$** (mean ± SD) [2] | 19.4 ± 6.2 | 22.3 ± 7.2 | 0.03 |
| **$PANSS_{general}$** (mean ± SD) [2] | 37.4 ± 7.7 | 44.0 ± 8.5 | <0.001* |
| **$PANSS_{total}$** (mean ± SD) [2] | 75.4 ± 14.8 | 86.9 ± 15.9 | <0.001* |
| **Estimated total intracranial volume, $10^6$ $mm^3$** (mean ± SD) [2] | 1.494 ± 0.167 | 1.513 ± 0.160 | 0.56 |

***Supplementary Table S2:*** *Demographic and clinical characteristics of participants included in the secondary analyses (N=114). AMI, amisulpride. ARI, aripiprazole. SD, standard deviation. GAF, Global Assessment of Functioning. [1] Mann-Whitney U-test, [2] two-sample t-test, [3] chi-squared test, [4] Fisher's exact test. Significant p-values are marked with *.*

| | FEP (N=61) | HC (N=53) | P-value | AMI (N=24) | ARI (N=37) | P-value |
|---|---|---|---|---|---|---|
| **Baseline** | | | | | | |
| **Age, years** (mean ± SD) [1] | 24.1 ± 5.6 | 23.0 ± 4.1 | 0.43 | 25.7 ± 6.1 | 23.0 ± 5.1 | 0.03* |
| **Sex** (M:F) [3] | 30:31 | 26:27 | 1.00 | 13:11 | 17:20 | 0.71 |
| **Cohort** (AMI:ARI) [3] | 24:37 | 25:28 | 0.51 | 24:0 | 0:37 | |
| **Handedness** (right: ambidextrous:left) [4] | 53:1:6 | 46:0:5 | 1.00 | 20:0:4 | 33:1:2 | 0.28 |
| **Handedness score, -100:100** (mean ± SD) [1] | 74.0 ± 47.7 | 79.3 ± 40.9 | 0.74 | 73.7 ± 50.3 | 74.2 ± 46.6 | 0.23 |
| **Parental socioeconomic status** (high:moderate:low) [4] | 15:36:8 | 8:18:3 | 0.90 | 5:15:2 | 10:21:6 | 0.74 |
| **Educational level** (higher education/self-employed:medium education:uneducated:student) [4] | 1:6:13:35 | 1:1:2:25 | 0.09 | 0:3:3:12 | 1:3:10:23 | 0.71 |
| **Years of education** (mean ± SD)[1] | 12.1 ± 2.2 | 13.4 ± 1.8 | 0.003* | 12.8 ± 2.4 | 11.6 ± 2.1 | 0.07 |
| **IQ** (mean ± SD) [2] | 99.4 ± 16.3 | 110.3 ± 12.1 | <0.001* | 101.7 ± 16.6 | 97.9 ± 16.1 | 0.40 |
| **Height, cm** (mean ± SD) [2] | 173.1 ± 9.7 | 174.4 ± 9.9 | 0.50 | 174.5 ± 9.6 | 172.2 ± 9.8 | 0.37 |
| **Weight, kg** (mean ± SD) [1] | 72.1 ± 15.2 | 68.7 ± 9.6 | 0.32 | 75.7 ± 17.1 | 69.8 ± 13.5 | 0.30 |
| **Alcohol use** (never:few times:regularly:harmful use: dependency) [4] | 3:15:39:4:- | 1:6:46:0:- | 0.02* | 2:2:18:2:- | 1:13:21:2:- | 0.07 |
| **Cannabis use** (never:few times:regularly:harmful use: dependency) [4] | 20:31:9:1:- | 21:29:3:0:- | 0.31 | 6:13:4:1:- | 14:18:5:0:- | 0.54 |
| **Benzodiazepines use** (never:few times:regularly:harmful use: dependency) [4] | 44:14:2:-:- | 52:0:0:-:- | <0.001* | 17:6:0:-:- | 27:8:2:-:- | 0.78 |
| **GAF, 0:100** (mean ± SD) [1] | 42.7 ± 11.3 | | | 40.5 ± 10.5 | 44.2 ± 11.7 | 0.18 |
| **$\text{PANSS}_{\text{positive}}$** (mean ± SD) [2] | 18.9 ± 4.5 | | | 19.8 ± 4.8 | 18.3 ± 4.3 | 0.19 |
| **$\text{PANSS}_{\text{negative}}$** (mean ± SD) [2] | 20.6 ± 5.8 | | | 21.3 ± 6.9 | 20.1 ± 5.0 | 0.43 |
| **$\text{PANSS}_{\text{general}}$** (mean ± SD) [2] | 38.7 ± 8.4 | | | 41.9 ± 9.9 | 36.6 ± 6.5 | 0.02* |
| **$\text{PANSS}_{\text{total}}$** (mean ± SD) [2] | 78.1 ± 15.8 | | | 83.0 ± 18.5 | 75.0 ± 13.2 | 0.05 |
| **Estimated total intracranial volume, $10^6$ $mm^3$** (mean ± SD) [2] | 1.489 ± 0.173 | 1.502 ± 0.153 | 0.67 | 1.490 ± 0.147 | 1.488 ± 0.190 | 0.96 |

***Supplementary Table S3:*** *Results of Welch's t-tests on baseline Z-scores from patients (N=86) and HCs (N=65). All IDPs showed were initially significant (p<0.05), although none remained significant after correction for multiple testing.*

| IDP | t-statistic | P-value | P-value corrected |
|---|---|---|---|
| Z *higher* in the FEP group | | | |
| Left-Inf-Lat-Vent | 2.034725 | 0.043763 | 0.547803 |
| rh_G_temp_sup-G_T_transv_thickness | 1.994144 | 0.048203 | 0.547803 |
| Z *lower* in FEP group | | | |
| Left-Hippocampus | -3.04172 | 0.002785 | 0.384555 |
| Right-Hippocampus | -2.91275 | 0.004135 | 0.384555 |
| Right-Cerebellum-Cortex | -2.61005 | 0.010013 | 0.547803 |
| Left-Cerebellum-Cortex | -2.38259 | 0.018474 | 0.547803 |
| Left-Accumbens-area | -2.32129 | 0.02164 | 0.547803 |
| lh_S_cingul-Marginalis_thickness | -2.2408 | 0.026603 | 0.547803 |
| lh_G_oc-temp_med-Parahip_thickness | -2.16309 | 0.032243 | 0.547803 |
| rh_G_oc-temp_med-Parahip_thickness | -2.10603 | 0.036917 | 0.547803 |
| SubCortGrayVol | -2.10174 | 0.037259 | 0.547803 |
| lh_S_postcentral_thickness | -2.08271 | 0.03911 | 0.547803 |
| lh_S_collat_transv_ant_thickness | -2.02328 | 0.045035 | 0.547803 |

***Supplementary Table S4:*** *Results of Wilcoxon signed-rank tests on $Z_{diff}$ scores for patients treated with* *amisulpride* *from baseline to six-week follow-up (N=41). All results displayed were significant after correction for multiple testing ($p_{corrected}$<0.05).*

| **IDP** | **W-statistic** | **P-value** | **P-value corrected** | **Mean** |
|---|---|---|---|---|
| *Increased* IDPs | | | | |
| Left-Caudate | 136 | 6.03646E-05 | 0.00780703 | 0.763775146 |
| Right-Caudate | 141 | 8.39466E-05 | 0.00780703 | 0.986049893 |
| Right-Putamen | 151 | 0.00015812 | 0.009803453 | 0.656149607 |

***Supplementary Table S5:*** *Results of Wilcoxon signed-rank tests on $Z_{diff}$ scores for patients treated with* *aripiprazole* *from baseline to six-week follow-up (N=45). All results displayed were significant after correction for multiple testing ($p_{corrected}$<0.05).*

| IDP | W-statistic | P-value | P-value corrected | Mean |
|---|---|---|---|---|
| *Increased* IDPs | | | | |
| Left-Caudate | 133 | 3.51365E-06 | 0.000653538 | 0.709677876 |
| Right-Caudate | 238 | 0.001230652 | 0.032700183 | 0.540567431 |
| *Decreased* IDPs | | | | |
| lh_G_front_inf-Orbital_thickness | 212 | 0.000368513 | 0.023215422 | -0.566958902 |
| rh_G_pariet_inf-Angular_thickness | 217 | 0.000469678 | 0.023215422 | -0.500507932 |
| lh_S_temporal_sup_thickness | 223 | 0.00062407 | 0.023215422 | -0.558940394 |
| lh_S_temporal_transverse_thickness | 223 | 0.00062407 | 0.023215422 | -0.517007048 |
| lh_S_precentral-inf-part_thickness | 235 | 0.001078144 | 0.032700183 | -0.402413126 |
| lh_G_temporal_middle_thickness | 244 | 0.001595285 | 0.033190017 | -0.583471045 |
| lh_G_cingul-Post-ventral_thickness | 248 | 0.00188957 | 0.033190017 | -0.486287422 |
| lh_S_subparietal_thickness | 250 | 0.002054234 | 0.033190017 | -0.538955756 |
| lh_G_cingul-Post-dorsal_thickness | 251 | 0.002141291 | 0.033190017 | -0.441077452 |
| rh_G_oc-temp_lat-fusifor_thickness | 251 | 0.002141291 | 0.033190017 | -0.549772352 |
| rh_S_temporal_sup_thickness | 257 | 0.002736544 | 0.039153631 | -0.60041231 |

***Supplementary Table S6:*** *Results of Wilcoxon signed-rank tests on patient $Z_{diff}$ scores from baseline to 6-month follow-up (N=46). All results displayed were significant after correction for multiple testing ($p_{corrected}$<0.05).*

| IDP | W-statistic | P-value | P-value corrected | Mean |
|---|---|---|---|---|
| *Increased* IDPs | | | | |
| Left-Caudate | 247 | 0.001011906 | 0.024300675 | 0.081015306 |
| Right-Pallidum | 253 | 0.001306488 | 0.024300675 | 0.176731527 |
| *Decreased* IDPs | | | | |
| lh_G&S_cingul-Mid-Ant_thickness | 160 | 1.08966E-05 | 0.001982837 | -0.709808833 |
| lh_G_front_sup_thickness | 171 | 2.13208E-05 | 0.001982837 | -0.344689878 |
| rh_S_precentral-inf-part_thickness | 216 | 0.000242981 | 0.015064838 | -0.44595754 |
| lh_G_precentral_thickness | 233 | 0.000543514 | 0.021162138 | -0.363526611 |
| rh_G&S_subcentral_thickness | 234 | 0.000568875 | 0.021162138 | -0.602003325 |
| rh_G_front_inf-Opercular_thickness | 239 | 0.000712557 | 0.022089265 | -0.499120289 |
| rh_G_temp_sup-Lateral_thickness | 250 | 0.001150717 | 0.024300675 | -0.352578934 |
| lh_S_collat_transv_post_thickness | 251 | 0.00120067 | 0.024300675 | -0.30592791 |
| lh_S_precentral-sup-part_thickness | 256 | 0.001481015 | 0.025042615 | -0.123214931 |
| lh_S_precentral-inf-part_thickness | 270 | 0.002605431 | 0.03727771 | -0.38229563 |
| rh_Lat_Fis-post_thickness | 270 | 0.002605431 | 0.03727771 | -0.31010438 |

***Supplementary Table S7:*** *Results of Wilcoxon signed-rank tests on patient $Z_{diff}$ scores from baseline to 1-year follow-up (N=15). All results displayed were significant after correction for multiple testing ($p_{corrected}<0.05$). Left cerebellum white matter was also increased in the HCs, and the left ACC and right opercular inferior frontal gyrus were also decreased in HCs (see Supplementary Table S9).*

| IDP | W-statistic | P-value | P-value corrected | Mean |
|---|---|---|---|---|
| *Increased* IDPs | | | | |
| lh_G_oc-temp_med-Parahip_thickness | 11 | 0.003356934 | 0.027147376 | 0.648108101 |
| lh_S_oc-temp_lat_thickness | 11 | 0.003356934 | 0.027147376 | 0.652013233 |
| Left-Cerebellum-White-Matter | 12 | 0.004272461 | 0.030564528 | 1.110829313 |
| lh_S_collat_transv_ant_thickness | 15 | 0.008361816 | 0.042035077 | 0.657444113 |
| Left-choroid-plexus | 16 | 0.010253906 | 0.047680664 | 0.837230382 |
| *Decreased* IDPs | | | | |
| lh_G_rectus_thickness | 0 | 6.10352E-05 | 0.003243583 | -1.288497762 |
| rh_G&S_transv_frontopol_thickness | 0 | 6.10352E-05 | 0.003243583 | -1.102141506 |
| lh_G&S_cingul-Ant_thickness | 1 | 0.00012207 | 0.003243583 | -1.469858252 |
| lh_G_front_sup_thickness | 1 | 0.00012207 | 0.003243583 | -1.565580951 |
| lh_S_suborbital_thickness | 1 | 0.00012207 | 0.003243583 | -1.203100464 |
| rh_S_front_middle_thickness | 1 | 0.00012207 | 0.003243583 | -1.058052873 |
| rh_S_orbital_lateral_thickness | 1 | 0.00012207 | 0.003243583 | -1.000112394 |
| rh_G_front_middle_thickness | 2 | 0.000183105 | 0.004257202 | -1.330094029 |
| lh_S_front_sup_thickness | 3 | 0.000305176 | 0.006306966 | -1.017212496 |
| lh_S_front_middle_thickness | 5 | 0.000610352 | 0.01032049 | -1.266711158 |
| lh_S_precentral-sup-part_thickness | 5 | 0.000610352 | 0.01032049 | -0.906021943 |
| rh_G&S_frontomargin_thickness | 6 | 0.000854492 | 0.013244629 | -0.860319641 |
| lh_G&S_transv_frontopol_thickness | 7 | 0.001159668 | 0.01348114 | -1.53585211 |
| lh_G_front_middle_thickness | 7 | 0.001159668 | 0.01348114 | -1.49339798 |
| lh_G_postcentral_thickness | 7 | 0.001159668 | 0.01348114 | -0.851088403 |
| rh_G_postcentral_thickness | 7 | 0.001159668 | 0.01348114 | -0.613619755 |
| lh_G_parietal_sup_thickness | 9 | 0.00201416 | 0.020812988 | -0.705166808 |
| rh_Lat_Fis-ant-Vertical_thickness | 9 | 0.00201416 | 0.020812988 | -1.875765661 |
| lh_S_circular_insula_ant_thickness | 10 | 0.002624512 | 0.024407959 | -1.074449536 |
| rh_Lat_Fis-ant-Horizont_thickness | 10 | 0.002624512 | 0.024407959 | -0.891195537 |
| rh_G_front_sup_thickness | 11 | 0.003356934 | 0.027147376 | -0.484005174 |
| lh_G&S_cingul-Mid-Ant_thickness | 12 | 0.004272461 | 0.030564528 | -0.964512785 |
| lh_S_precentral-inf-part_thickness | 12 | 0.004272461 | 0.030564528 | -0.756316913 |
| lh_S_orbital-H_Shaped_thickness | 13 | 0.005371094 | 0.034449084 | -0.596616846 |
| rh_G_front_inf-Opercular_thickness | 13 | 0.005371094 | 0.034449084 | -0.893914052 |
| rh_Pole_temporal_thickness | 13 | 0.005371094 | 0.034449084 | -0.519066752 |
| lh_G_occipital_sup_thickness | 14 | 0.006713867 | 0.036728803 | -0.806475905 |
| lh_S_front_inf_thickness | 14 | 0.006713867 | 0.036728803 | -0.854580052 |
| lh_MeanThickness_thickness | 14 | 0.006713867 | 0.036728803 | -0.656234223 |
| rh_S_circular_insula_inf_thickness | 14 | 0.006713867 | 0.036728803 | -0.679166137 |
| rh_S_front_inf_thickness | 14 | 0.006713867 | 0.036728803 | -0.950848659 |
| lh_G&S_frontomargin_thickness | 15 | 0.008361816 | 0.042035077 | -1.357399225 |
| rh_S_front_sup_thickness | 15 | 0.008361816 | 0.042035077 | -0.661302722 |
| lh_Pole_occipital_thickness | 16 | 0.010253906 | 0.047680664 | -0.79154453 |
| rh_G&S_subcentral_thickness | 16 | 0.010253906 | 0.047680664 | -0.865480958 |

***Supplementary Table S8:*** *Results of Wilcoxon signed-rank tests on patient $Z_{diff}$ scores from baseline to 2-year follow-up (N=36). All results displayed were significant after correction for multiple testing ($p_{corrected}$<0.05). Left cerebellum white matter was also increased in HCs (see Supplementary Table S9).*

| IDP | W-statistic | P-value | P-value corrected | Mean |
|---|---|---|---|---|
| *Increased* IDPs | | | | |
| Right-Cerebellum-White-Matter | 69 | 7.36583E-06 | 0.001370044 | 0.460334256 |
| Left-Cerebellum-White-Matter* | 90 | 5.14205E-05 | 0.003188071 | 0.436115284 |
| *Decreased* IDPs | | | | |
| lh_G&S_cingul-Mid-Ant_thickness | 78 | 1.76393E-05 | 0.001640453 | -0.949496326 |
| lh_Lat_Fis-ant-Horizont_thickness | 112 | 0.000289781 | 0.011588735 | -0.559652556 |
| lh_S_precentral-sup-part_thickness | 113 | 0.000311525 | 0.011588735 | -0.154751337 |
| lh_G&S_cingul-Ant_thickness | 116 | 0.000385896 | 0.011962772 | -0.746524017 |
| lh_S_pericallosal_thickness | 128 | 0.000870808 | 0.021603714 | -0.7197586 |
| rh_S_precentral-inf-part_thickness | 129 | 0.000929192 | 0.021603714 | -0.587737577 |
| lh_G_front_sup_thickness | 138 | 0.00163483 | 0.033786488 | -0.302501651 |
| rh_Lat_Fis-ant-Horizont_thickness | 146 | 0.002628042 | 0.04707246 | -0.359358511 |
| lh_G&S_cingul-Mid-Post_thickness | 147 | 0.002783855 | 0.04707246 | -0.520772251 |

***Supplementary Table S9:*** *Results of Wilcoxon signed-rank tests on HC $Z_{diff}$ scores from baseline to 1-year and 2-year follow-ups (N=16 and N=35, respectively). All results displayed are significant after correction for multiple testing ($p_{corrected}$<0.05).*

| IDP | W-statistic | P-value | P-value corrected | Mean |
|---|---|---|---|---|
| **BASELINE TO 1-YEAR FOLLOW-UP** | | | | |
| *Increased* IDPs | | | | |
| Left-Cerebellum-White-Matter | 5 | 0.000305 | 0.028381 | 0.928712 |
| rh_S_cingul-Marginalis_thickness | 7 | 0.00058 | 0.03595 | 0.967961 |
| rh_S_subparietal_thickness | 10 | 0.001312 | 0.049667 | 0.829751 |
| rh_G_precuneus_thickness | 11 | 0.001678 | 0.049667 | 0.940739 |
| rh_S_parieto_occipital_thickness | 12 | 0.002136 | 0.049667 | 0.903804 |
| *Decreased* IDPs | | | | |
| rh_G_front_inf-Triangul_thickness | 5 | 0.000305 | 0.028381 | -1.14751 |
| lh_G&S_cingul-Ant_thickness | 11 | 0.001678 | 0.049667 | -1.0307 |
| rh_G_front_inf-Opercular_thickness | 12 | 0.002136 | 0.049667 | -0.91317 |
| **BASELINE TO 2-YEAR FOLLOW-UP** | | | | |
| *Increased IDPs* | | | | |
| Left-Cerebellum-White-Matter | 92 | 0.000118118 | 0.020905991 | 0.262937392 |
| rh_G_precuneus_thickness | 100 | 0.000224796 | 0.020905991 | 0.69193347 |
| rh_G_Ins_lg&S_cent_ins_thickness | 113 | 0.00059151 | 0.036673597 | 0.400542926 |

### References

1. Rutherford S, Fraza C, Dinga R, Kia SM, Wolfers T, Zabihi M, m.fl. Charting brain growth and aging at high spatial precision. eLife. 1. februar 2022;11:e72904.

2. Fraza CJ, Dinga R, Beckmann CF, Marquand AF. Warped Bayesian linear regression for normative modelling of big data. NeuroImage. december 2021;245:118715.

3. Bučková BR, Fraza C, Rehák R, Kolenič M, Beckmann CF, Španiel F, m.fl. Using normative models pre-trained on cross-sectional data to evaluate intra-individual longitudinal changes in neuroimaging data. eLife. 12. marts 2025;13:RP95823.
